# Association between *Clostridioides difficile* infection and colorectal cancer incidence and mortality in a National Veterans Affairs Cohort

**DOI:** 10.64898/2026.05.20.26353667

**Authors:** SB Rifkin, NO Markham, SM Anderson, O Wilson, MJ Shrubsole, CL Sears, K Rao

## Abstract

**Background:** Recent mouse model data demonstrate that chronic colonization with toxigenic *Clostridioides difficile* promotes colonic tumorigenesis via intraluminal toxin B (TcdB), its main virulence factor. In a prior multisite hospital cohort, we found that history of positive *tcdB* stool testing was associated with increased CRC risk in a dose-dependent manner, though limited by small sample size. We aimed to validate this association in a larger cohort with extended follow-up and greater geographic distribution using the Veterans Health Administration (VHA) Corporate Data Warehouse (CDW).

**Methods:** We conducted a retrospective cohort study among adults receiving care through the VA from 2000-2025 who underwent *C. difficile* testing. Data collected from the VHA CDW and National Death Index (NDI) included demographics, comorbidities, medications, CRC risk factors, and cancer incidence and death. The first *C. difficile* test date defined cohort entry; individuals with prior CRC were excluded. Ever *C. difficile* positivity was defined by a positive PCR or EIA results. The number of positive tests (episodes) was also determined to define recurrent positivity. Follow-up time ended at the first occurrence of CRC incidence or mortality, death from other causes, or censor date. Follow-up time was split for individuals who converted from negative to positive, with follow-up time updated accordingly. Multivariable Cox proportional hazards models were used to estimate hazard ratios (HRs) for *C. difficile* exposure and CRC incidence and mortality after adjustment for confounders. Tests for linear trend and tests for interaction were conducted to assess effect modification by sex and IBD status, while time-lag intervals were evaluated for 1, 3, 5, and 10 years before the outcome.

**Results:** Among 806,844 veterans with *C. difficile testing*, those with positive tests were more likely to be older, male, to have diabetes, to use aspirin, and to have a lower BMI than those with negative tests. Race and IBD prevalence were similar between the groups. There was no overall association between ever *C. difficile* positivity and CRC incidence (HR = 0.99, 95% CI 0.93-1.05). However, recurrent *C. difficile* positivity was associated with increased risk in a dose-response manner [2-3 episodes HR = 1.30 (95% CI 1.16-1.47), and >3 episodes HR = 1.58 (95% CI 1.17-2.14) compared to negative tests; ptrend< 0.001]. Further, ever *C. difficile* positivity was associated with increased CRC mortality risk (HR = 1.21, 95% CI 1.13-1.30; *p* < 0.001). Recurrent *C. difficile* positivity was associated with increased mortality risk but was particularly strong for those with >3 episodes among individuals with IBD (HR=3.84, 95% CI 1.98-7.45). In sensitivity analyses, the increased risk of CRC incidence and mortality attenuated beyond 10 years.

**Conclusion:** Prior positive *C. difficile* testing was associated with increased CRC incidence and mortality in a dose-dependent manner, particularly among patients with IBD. These findings extend animal model evidence, epidemiologically establishing *C. difficile* presence as an independent risk factor for subsequent colorectal tumorigenesis and supporting investigation into recurrent CDI, especially among patients with IBD, as a potential modifiable CRC risk factor.

## Introduction

Sporadic colorectal cancer (CRC) remains the third leading cause of cancer incidence and second leading cause of cancer mortality worldwide (1). Increasing evidence suggests that gut microbiome alterations as well as specific enteric bacteria may contribute to colorectal carcinogenesis (2). The microbiome both influences and is influenced by host risk factors including diet, tobacco use, alcohol consumption, and obesity, all of which are established CRC risk factors (3). These bidirectional relationships form a complex web that remains inadequately studied.

*Clostridioides difficile* is an increasingly common enteric pathogen, with asymptomatic carriage estimated to occur in ∼10% of the general population (4) and higher rates in infants and hospitalized patients. Clinically significant *C. difficile* infection (CDI) occurs after germination of colonizing *C. difficile* spores and toxin production resulting in a broad spectrum of disease ranging from mild diarrheal illness to fulminant and life-threatening colitis and sepsis. Over the past 30 years, the epidemiology of CDI has changed significantly with a dramatic increase in incidence and recurrence (rCDI). CDI occurs in over 500,000 people per year in the United States alone (5) with approximately 25% experiencing a recurrent infection. After an initial rCDI episode, the probability of rCDI increases with each subsequent rCDI event.

As the burden of CDI grows, the long-term sequelae of infection are increasingly recognized. Notably, *C. difficile* colonization may persist following treatment with some patients shedding spores for at least 5-6 months after initial treatment of CDI, potentially leading to prolonged host exposure to *C. difficile* cytotoxins and other virulence factors (6). Emerging evidence suggests that CDI is associated with lasting microbiome disruption, chronic gastrointestinal symptoms, inflammatory conditions, increased mortality, and potentially CRC (7-12). An experimental heterozygous <u>A</u>denomatous <u>p</u>olyposis <u>c</u>oli (*Apc*^*+/-*^) multiple intestinal neoplasia (Min) mouse model demonstrated that chronic colonization with toxigenic *C. difficile* promoted colonic tumorigenesis via production of intraluminal toxin B (TcdB), its main virulence factor. TcdB activated protumorigenic inflammatory pathways and prooncogenic Wnt signaling to effect these changes (10).

Prior epidemiological studies evaluating the association between *C. difficile* exposure and CRC risk have found conflicting results (13-15). The limited prior investigations were based on administrative billing codes to define CDI, raising concerns of exposure misclassification, and lacked temporal separation between CDI diagnosis and CRC development. Few studies assessing CRC risk have used laboratory diagnosis to define *C. difficile* exposure or have evaluated recurrent/persistent C. *difficile* positivity or dose-dependent effects. In a prior multisite hospital-based cohort, we found that history of positive *C. difficile* stool testing was associated with increased CRC risk in a dose-dependent manner, though it was limited by small sample size and short follow-up time, impacting our ability to evaluate subgroup analyses (12).

To address these limitations and validate our previous findings, we performed a retrospective cohort study using the Veterans Health Administration (VHA) Corporate Data Warehouse, which provides a large, national cohort with extended longitudinal follow-up. We evaluated CRC incidence and mortality in individuals with and without CDI and examined whether rCDI is associated with a dose-dependent increase in CRC risk.

## Methods

We conducted a retrospective study of adults receiving healthcare through the Veterans Health Administration (VHA) from 2000 to 2025 who underwent testing for *Clostridioides difficile* infection (CDI). Data were obtained from the VHA corporate data warehouse and the National Death Index (NDI), and included demographics, anthropometric measurements, medical diagnoses, microbiological test results, medication prescriptions, colon cancer risk factors, and laboratory results. The date of the first laboratory test for *C. difficile* served as the cohort entry (index) date. Individuals with a prior history of CRC or total colectomy before cohort entry were excluded. IRB approval was obtained from the Nashville VHA.

### Exposure

The primary exposures of interest were a history of *C. difficile* test positivity and a cumulative number of positive *C. difficile* tests occurring between cohort entry and CRC diagnosis or end of follow-up. *C. difficile* test positivity was defined by a positive result on either the *C. difficile* Toxin B gene (*tcdB)* polymerase chain reaction (PCR) or *C. difficile* Toxin B protein (TcdB) enzyme immunoassay (EIA) test. Individuals were classified as *C. difficile* exposed if any *C. difficile* test performed on or after the index date and before CRC diagnosis or study end was positive. Those with exclusively negative tests were classified as unexposed. For individuals who initially tested negative but subsequently tested positive, exposure status was treated as time-varying, with follow-up time divided into unexposed (prior to first positive test) and exposed (after the first positive test) periods.

Participants with any exposed follow-up time were categorized using both binary (never vs. ever exposed) and nominal definition based on cumulative number of positive *C. difficile* tests: no positive tests (CD=0), one positive test (CD=1), two to three positive tests spaced more than 30 days apart (CD >1 & <4), or more than three positive tests (CD >3). In the nominal analysis, individuals could contribute time to multiple exposure categories as additional positive tests occurred. This approach was used to assess potential dose-response effects suggested by prior data.

### Covariates

Potential confounders were selected *a priori* based on established risk factors for CDI and CRC, including age, sex, race, body mass index (BMI), diabetes mellitus (DM), prior antibiotic use, aspirin use, history of inflammatory bowel disease (IBD), tobacco use and frailty all assessed at cohort entry. Medical comorbidities were identified using ICD-9 and ICD-10 codes recorded in the year prior to cohort entry. BMI was calculated from available anthropometric data. Prior aspirin use was ascertained from medication records documenting any aspirin prescribed within the year prior to cohort entry. Prior antibiotic use was identified from medication records for all systemic antibiotics except oral vancomycin or fidaxomicin (as these are treatments for CDI) within 90 days before cohort entry. Tobacco status (never, former, or current use) was extracted from medication records. Frailty was measured using the Veterans Affairs Frailty Index (VAFI), a validated claims-based measure of biological age and comorbidity burden, calculated using ICD-9 and ICD-10 diagnoses codes.

### Outcomes

The primary outcomes were incident CRC, ascertained using the VHA Central Cancer Registry and Oncology domain and supplemented by CRC-related deaths from the NDI capture cases not recorded in registry data (16, 17). CRC diagnoses were further categorized by tumor location (right colon, left colon, or rectum) using ICD-9 and ICD-10 codes. The secondary outcome was CRC mortality, defined separately as CRC-related deaths based on NDI data alone.

### Statistical Analysis

Baseline characteristics were compared between individuals with and without *C. difficile* positive testing using Student’s t test and chi-square tests for continuous and categorical variables, respectively. Time to event analyses were performed to evaluate the association of prior *C. difficile* positive exposure with risk of incident CRC. Participants were followed from the date of their index *C. difficile* test until they were diagnosed with CRC, death, or censoring (last encounter date in the system), whichever occurred first.

Unadjusted and multivariable Cox proportional analysis were used to estimate the hazard ratios (HRs) and 95% confidence intervals (CI) for the association between *C. difficile* exposure and incident CRC overall, as well as by tumor site (right colon, left colon, and rectum). Risk factors, microbiome composition, and tumor biology differ between the right, left colon and rectum, which is why we evaluated outcome by anatomical location. Exposure was modeled as both a binary variable (ever vs. never) and as a categorical variable (based on cumulative number of positive tests). Covariates included in the final model were selected *a priori* based on established epidemiologic CRC risk factors (3). The final multivariable Cox model included age, sex, diabetes mellitus (yes/no), BMI category (<18, 18-25, 25-30, >30), aspirin use (yes/no), tobacco use, race (white, black, Asian, Native Hawaiian/Pacific Islander, American Indian), frailty and personal history of IBD status (yes/no).

This final multivariable model was also studied across strata of sex and IBD, given known sex differences in CRC epidemiology and increased risk of CDI and inflammation-driven CRC risk in IBD. Effect modification was investigated by including interaction terms between *C. difficile* exposure and covariates of interest and applying a likelihood ratio test with p values less than 0.05 considered statistically significant. Lagged exposure analyses were conducted excluding CRC outcomes occurring 1, 3, 5, and 10 years after the index date. Proportional hazards assumptions were tested by including an interaction term between *C. difficile* exposure and follow-up time in a Cox regression model. There was no evidence that proportional hazard assumptions were violated. Secondary analyses evaluated the association between *C. difficile* exposure and CRC mortality, using the same multivariable modeling strategy as in the primary analyses. All analyses were conducted using R (version 4.4.2), with the survival package (version 3.8.3).

## Results

Among 806,844 veterans who underwent testing for CDI from 2000 to 2025, individuals with more *C. difficile* exposures were more likely to be older (mean age 64 vs 67-68), more frail (0.22 vs 0.25-0.26), more often male (92% vs 94-95%), have higher prevalences of DM (36% vs 38-41%), IBD (13% vs 14-17%), and aspirin use (47 vs 52-55%, p < 0.001) compared to those with none or a single episode, while race distribution was similar across number of *C. difficile* exposures (see Table 1).

**Table 1.** Baseline characteristics of patients according to number of *C. difficile* infections (CDI), VA (2000-2025)

| Characteristic | CDI=0<br>N = 652,860 | CDI=1<br>N = 116,101 | CDI=2-3<br>N = 32,511 | CDI ≥3<br>N = 5,372 | p-value |
| --- | --- | --- | --- | --- | --- |
| Age, years | 64 (15) | 67 (14) | 68 (13) | 66 (14) | <0.001 |
| Sex |  |  |  |  | <0.001 |
| Female | 52,983 (8.1) | 7,138 (6.1) | 1,829 (5.6) | 291 (5.4) |  |
| Male | 599,877 (92) | 108,963 (94) | 30,682 (94) | 5,081 (95) |  |
| BMI | 29 (6) | 28 (6) | 28 (6) | 28 (6) | <0.001 |
| Diabetes mellitus | 233,871 (36) | 43,711 (38) | 13,195 (41) | 2,222 (41) | <0.001 |
| Aspirin use | 304,547 (47) | 59,933 (52) | 17,843 (55) | 2,925 (54) | <0.001 |
| Smoking status |  |  |  |  | <0.001 |
| Never | 189,361 (29) | 31,628 (27) | 8,898 (27) | 1,454 (27) |  |
| Current | 203,638 (31) | 35,627 (31) | 9,633 (30) | 1,575 (29) |  |
| Former | 143,720 (22) | 25,920 (22) | 7,704 (24) | 1,280 (24) |  |
| Unknown | 116,141 (18) | 22,926 (20) | 6,276 (19) | 1,063 (20) |  |
| Inflammatory bowel disease | 86,874 (13) | 14,810 (13) | 4,512 (14) | 887 (17) | <0.001 |
| Frailty index | 0.22 (0.14) | 0.25 (0.14) | 0.26 (0.14) | 0.26 (0.14) | <0.001 |
| Race |  |  |  |  | <0.001 |
| American Indian or Alaska Native | 4,274 (0.7) | 706 (0.6) | 202 (0.6) | 31 (0.6) |  |
| Asian | 3,785 (0.6) | 503 (0.4) | 121 (0.4) | 22 (0.4) |  |
| Black or African American | 103,869 (16) | 18,024 (16) | 4,911 (15) | 856 (16) |  |
| Native Hawaiian or Other Pacific Islander | 4,306 (0.7) | 704 (0.6) | 201 (0.6) | 31 (0.6) |  |
| No matching concept | 63,728 (9.8) | 13,681 (12) | 3,443 (11) | 470 (8.7) |  |
| White | 472,898 (72) | 82,483 (71) | 23,633 (73) | 3,962 (74) |  |
| Follow up time | 2,012 (1,985) | 1,771 (1,951) | 1,867 (1,857) | 2,311 (1,915) | <0.001 |

1. Values are mean (SD) or n (%). 2. p-values from one-way analysis of means (Welch ANOVA) for continuous variables and Pearson's $\chi^2$ test for categorical variables.

The cohort contributed 7,912,994 total person-years of follow-up. At baseline, the median age of the cohort was 66 years, 16% were Black, and 8% were female. During the follow-up time, there were 9,758 incident CRC cases: 1363 (16%) of these incident CRC cases were recorded in individuals with any *C. difficile* exposure. The overall CRC incidence rate per 1,000 patient-years was 1.22, 0.82, 0.55, and 0.29 in individuals with no, one, 2-3, and >3 positive *C. difficile* tests during the follow-up window, respectively.

Ever *C. difficile* test positivity was not associated with overall risk of CRC (HR = 0.99, 95% CI 0.93-1.05). While an increased risk was observed among females, this association did not reach statistical significance (HR 1.32, 95% CI 0.97-1.82), and no association was seen among males. However, recurrent *C. difficile* test positivity was associated with increased risk of CRC incidence. This analysis showed a dose-response pattern, one episode HR = 0.92 (95% CI 0.86-0.98), 2-3 episodes HR = 1.30 (95% CI 1.16-1.47), and >3 episodes HR = 1.58 (95% CI 1.17-2.14), in comparison to no episodes. In the stratified analysis among men, the same dose response pattern was seen. In women, no clear dose-response was seen; however there was a non-significant increased risk among women with more than one episode (HR= 1.33 (95% CI 0.95-1.87). When sex was included in the full model the interaction term between *C. difficile* exposure and sex was borderline significant (p=0.06). Notably, women were underrepresented in this dataset of veterans, resulting in fewer women in the category of increased episodes of *C. difficile* test positivity (see Table 2, Figure 1).

**Table 2.** Associations of Any CDI and Dose-Dependent *Clostridioides difficile* Exposure With Colorectal Cancer Incidence by Sex and IBD Status (VHA, 2000–2025)

|  | CDI Status |  |  |  |  | CDI Exposures |  |  |  |  |  |  |  |  |  |
| --- | --- | --- | --- | --- | --- | --- | --- | --- | --- | --- | --- | --- | --- | --- | --- |
|  | Never |  | Ever |  |  | CD=0 |  | CD = 1 |  | CD=2-3 |  | CD>3 |  | P trend | P interaction |
|  | Event s/py | HR (95% CI)<sup>1</sup> | Event s/py | HR (95% CI) |  | Event s/py | HR (95% CI)<sup>1</sup> | Event s/py | HR (95% CI)<sup>1</sup> | Events/py | HR (95% CI)<sup>1</sup> | Events/py | HR (95% CI)<sup>1</sup> |  |  |
| Overall | 7375/21915<br>42752 |  | 1363/69670<br>0121 | 0.99 (0.93-1.05) |  | 7375/21915<br>42752 |  | 1048/46295<br>9538 | 0.92 (0.86-0.98) | 273/180<br>918319 | 1.30 (1.16-1.47) | 42/528222<br>64 | 1.58 (1.17-2.14) | 0.003 |  |
| Female | 223/21066<br>373 |  | 47/6124039<br>2 | 1.32 (0.97-1.82) |  | 223/21066<br>373 |  | 39/4133671<br>5 | 1.33 (0.95-1.87) | 8/15786<br>461 | 1.48 (0.73-3.02) | 0/4117216 | 0.00 (0.00-Inf) | >0.9 | 0.05 |
| Male | 7152/19704<br>76379 |  | 1316/63545<br>9729 | 0.98 (0.93-1.04) |  | 7152/19704<br>76379 |  | 1009/42162<br>2823 | 0.91 (0.85-0.97) | 265/165131858 | 1.30 (1.15-1.47) | 42/487050<br>48 | 1.63 (1.20-2.21) | 0.002 |  |
| IBD | 932/414066<br>306 |  | 186/137376<br>460 | 1.18 (1.01-1.38) |  | 932/414066<br>306 |  | 139/875358<br>72 | 1.09 (0.91-1.31) | 39/12698 | 1.49 (1.08-2.06) | 8/3553 | 1.99 (0.99-3.99) | 0.005 | 0.2 |
| no IBD | 6443/17774<br>76446 |  | 1177/55932<br>3661 | 0.96 (0.91-1.03) |  | 6443/17774<br>76446 |  | 909/375423<br>666 | 0.90 (0.84-0.96) | 234/143984518 | 1.28 (1.12-1.45) | 34/399154<br>77 | 1.50 (1.07-2.11) | 0.3 |  |

<sup>1</sup>Hazard ratios (HRs) and 95% confidence intervals (CIs) were estimated using multivariable Cox proportional hazards regression adjusted for age, sex, body mass index (BMI) category, aspirin use, diabetes mellitus (DM), inflammatory bowel disease (IBD), frailty, and race.

**Figure 1.**
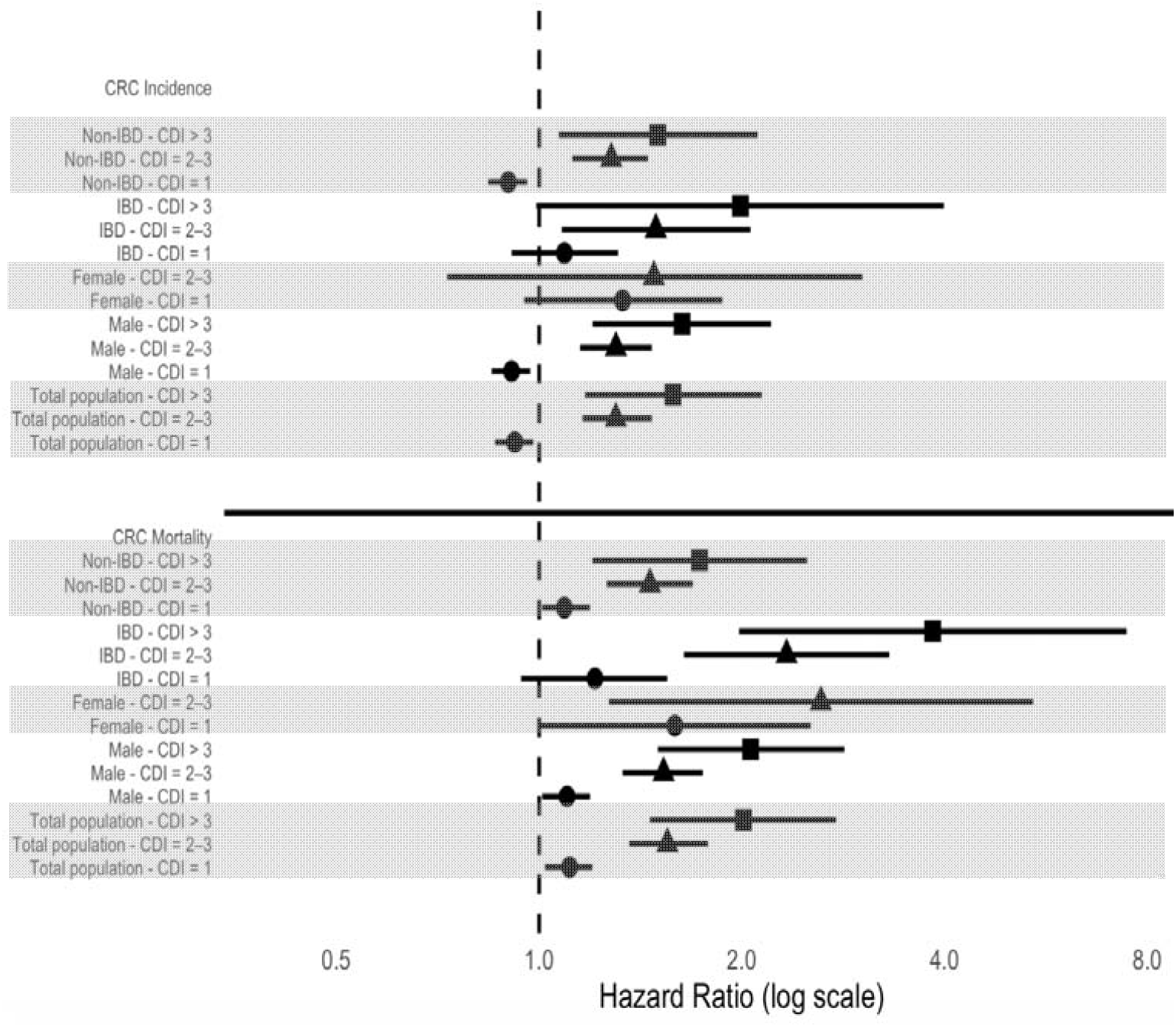
Forest plot of HR (95% CI) for the association between number of CDI and CRC outcomes by Nominal Dose Effect of CD Exposure (1, 2-3 and >3) in Comparison to No CD Exposure and Colorectal Cancer Incidence and Colorectal Cancer Mortality Risk, Overall and By Sex and IBD Status, VA (2000-2025)

In a stratified analysis by IBD status, ever *C. difficile* test positivity was associated with increased risk of CRC among patients with IBD (HR= 1.18, 95% CI, 1.01-1.38), but not among non-IBD patients (HR=0.96, 95% CI, 0.91-1.03). Similar dose-dependent increases in CRC risk were observed in both IBD and non-IBD patients with increasing number of episodes. The magnitude of associations was slighter stronger in the IBD group (2-3 episodes: HR=1.49 (1.08-2.06); >3 episodes: HR=1.99 (95% CI 0.99-3.99), vs. no positivity, ptrend = 0.005) compared with the non-IBD group (2-3 episodes: HR=1.28 (95% CI 1.12-1.45); >3 episodes: HR=1.50 (95% CI 1.07-2.11), ptrend=0.3) but the confidence intervals overlapped. Furthermore, there was no statistically significant interaction according to IBD status, when IBD was included in the full model, (pinteraction=0.2), (see Table 2, Figure 1).

Finally, a dose response relationship was observed for right colon cancers with HR 1.29 (95% CI 1.03-1.62) for 2-3 episodes and HR 1.87 (95% CI 1.14-3.06) for >3 episodes. A similar association was observed in the left colon with HR 1.59 (95% CI 1.20-2.11) for 2-3 episodes but no clear dose response relationship was seen for >3 episodes (HR 1.04, 95% CI 0.39-2.78). No significant association was observed for rectal cancer incidence (see Table 3).

**Table 3.**
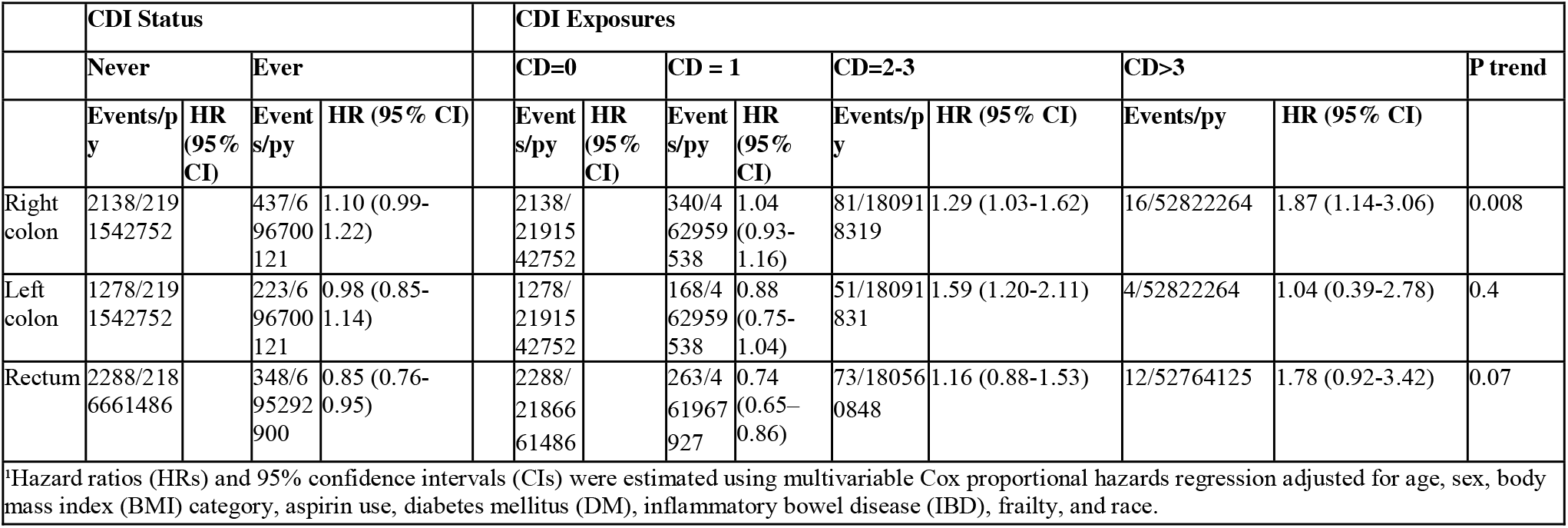
Associations of Any CDI and Dose-Dependent *Clostridioides difficile* Exposure With Colorectal Cancer Incidence by anatomical site (VHA, 2000–2025)

|  | CDI Status |  |  |  |  | CDI Exposures |  |  |  |  |  |  |  |  |
| --- | --- | --- | --- | --- | --- | --- | --- | --- | --- | --- | --- | --- | --- | --- |
|  | Never |  | Ever |  |  | CD=0 |  | CD = 1 |  | CD=2-3 |  | CD>3 |  | P trend |
|  | Events/p<br>y | HR<br>(95%<br>CI) | Event<br>s/py | HR (95% CI) |  | Event<br>s/py | HR<br>(95%<br>CI) | Event<br>s/py | HR<br>(95%<br>CI) | Events/p<br>y | HR (95% CI) | Events/py | HR (95% CI) |  |
| Right<br>colon | 2138/219<br>1542752 |  | 437/6<br>96700<br>121 | 1.10 (0.99-1.22) |  | 2138/<br>21915<br>42752 |  | 340/4<br>62959<br>538 | 1.04<br>(0.93-1.16) | 81/18091<br>8319 | 1.29 (1.03-1.62) | 16/52822264 | 1.87 (1.14-3.06) | 0.008 |
| Left<br>colon | 1278/219<br>1542752 |  | 223/6<br>96700<br>121 | 0.98 (0.85-1.14) |  | 1278/<br>21915<br>42752 |  | 168/4<br>62959<br>538 | 0.88<br>(0.75-1.04) | 51/18091<br>831 | 1.59 (1.20-2.11) | 4/52822264 | 1.04 (0.39-2.78) | 0.4 |
| Rectum | 2288/218<br>6661486 |  | 348/6<br>95292<br>900 | 0.85 (0.76-0.95) |  | 2288/<br>21866<br>61486 |  | 263/4<br>61967<br>927 | 0.74<br>(0.65–<br>0.86) | 73/18056<br>0848 | 1.16 (0.88-1.53) | 12/52764125 | 1.78 (0.92-3.42) | 0.07 |

\*Hazard ratios (HRs) and 95% confidence intervals (CIs) were estimated using multivariable Cox proportional hazards regression adjusted for age, sex, body mass index (BMI) category, aspirin use, diabetes mellitus (DM), inflammatory bowel disease (IBD), frailty, and race.

Remarkably, compared to CRC incidence where any *C. difficile* test positivity was not associated with overall risk, CRC mortality had a strong association with any *C. difficile* test positivity. Any number of positive *C. difficile* tests was associated with a significantly increased risk of CRC mortality overall (HR 1.21, 95% CI 1.13-1.30), with a pronounced risk among women (HR 1.72, 95% CI 1.14-2.59) and a significant but weaker association among men (HR 1.20, 95% CI 1.11-1.28). Furthermore, even a single positive test was associated with significantly increased risk, and a stronger dose-dependent relationship was observed for CRC mortality with increasing *C. difficile* test positivity exposures (1 episode: HR 1.11, 95% CI 1.02-1.20; 2-3 episodes: HR 1.55, 95% CI 1.36-1.78; >3 episodes: HR 2.01, 95% CI 1.46-2.76), compared to CRC incidence (see Figure 1, Table 5).

**Table 4.** Associations of Any CDI and Dose-Dependent *Clostridioides difficile* Exposure With Colorectal Cancer Incidence by time lag (VHA, 2000–2025)

|  | CDI Status |  |  |  |  | CDI Exposures |  |  |  |  |  |  |  |  |
| --- | --- | --- | --- | --- | --- | --- | --- | --- | --- | --- | --- | --- | --- | --- |
|  | Never |  | Ever |  |  | CD=0 |  | CD = 1 |  | CD=2-3 |  | CD>3 |  | P trend |
|  | Events/p<br>y | HR<br>(95%<br>CI) | Event<br>s/py | HR (95% CI) |  | Event<br>s/py | HR<br>(95%<br>CI) | Event<br>s/py | HR (95% CI) | Events/p<br>y | HR (95% CI) | Events/p<br>y | HR (95% CI) |  |
| 3-year<br>lag | 2537/204<br>8366068 |  | 574/6<br>47099<br>265 | 1.16 (1.06-1.27) |  | 2537/<br>20483<br>66068 |  | 412/4<br>28951<br>298 | 1.09 (0.98-1.21) | 132/1679<br>21998 | 1.32 (1.10-1.57) | 30/50225<br>969 | 1.73 (1.21-2.48) | <0.001 |
| 5-year<br>lag | 1722/187<br>0074835 |  | 397/5<br>86024<br>348 | 1.18 (1.06-1.32) |  | 1722/<br>18700<br>74835 |  | 284/3<br>88504<br>567 | 1.11 (0.98-1.26) | 95/15144<br>5094 | 1.41 (1.15-1.74) | 18/46074<br>687 | 1.12 (0.76-1.67) | <0.001 |
| 10-year<br>lag | 613/1224<br>943071 |  | 138/3<br>80391<br>378 | 1.13 (0.94-1.36) |  | 613/1<br>22494<br>3071 |  | 106/2<br>52573<br>399 | 1.12 (0.91-1.38) | 26/96564<br>759 | 1.49 (0.94-2.38) | 6/312532<br>20 | 1.29 (0.58-2.89) | 0.2 |

\*Hazard ratios (HRs) and 95% confidence intervals (CIs) were estimated using multivariable Cox proportional hazards regression adjusted for age, sex, body mass index (BMI) category, aspirin use, diabetes mellitus (DM), inflammatory bowel disease (IBD), frailty, and race.

**Table 5.**
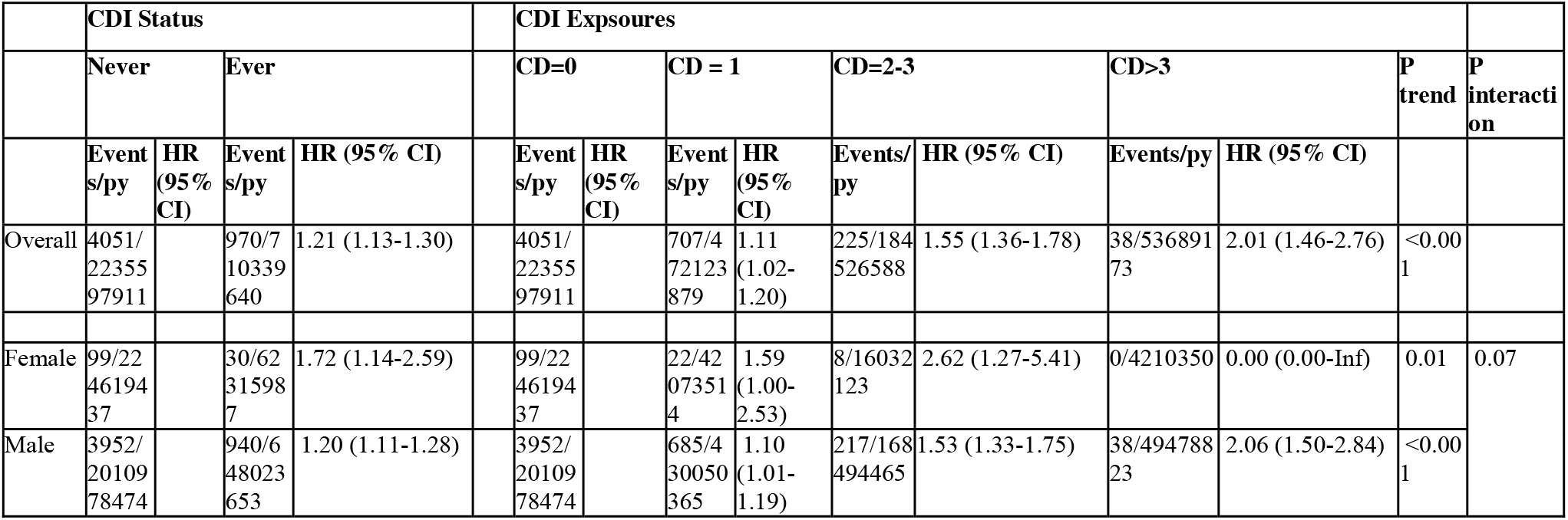
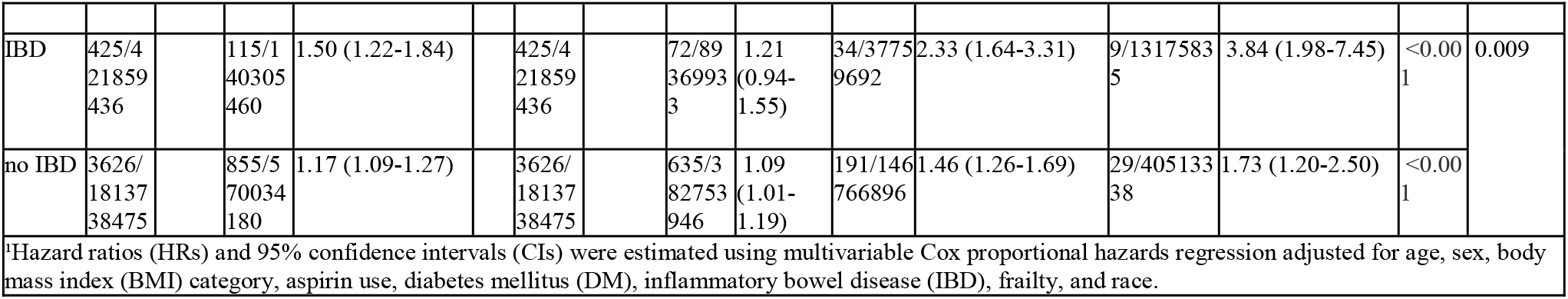
Associations of Any CDI and Dose-Dependent *Clostridioides difficile* Exposure With Colorectal Cancer Mortality by Sex and IBD Status (VHA, 2000–2025)

As with CRC incidence, this dose-dependent association between *C. difficile* positive exposure and CRC mortality was even more pronounced among patients with IBD, who had substantially higher mortality risks at higher doses (2-3 episodes HR = 2.33, 95% CI 1.64-3.31; >3 episodes: HR 3.84, 95% CI 1.98-7.45) compared to individuals without IBD. Additionally, the interaction term p-value for *C. difficile* exposure and IBD was significant (p=0.009) also consistent with effect modification by IBD (see Figure 1, Table 5).

Likewise, risk of mortality due to right-sided colon cancer increased progressively with recurrent *C. difficile* test positivity with HR 2.00 (95% CI 1.25-3.20) for one episode, HR 2.44 (95% CI 1.05-5.65) for 2-3 episodes, and HR 6.19 (95% CI 1.50-25.4) for > 3 episodes in comparison to no positivity. Similarly, rectal cancer mortality was significantly greater at higher exposure levels (HR 1.96 (95% CI 1.53-2.52) for 2-3 episodes and HR 2.36 (95% CI 1.27-4.41) for >3 episodes) but, unlike right colon cancer, was not increased after only one episode. No significant association between *C. difficile* test positivity and left colon cancer mortality was observed (see Table 6).

**Table 6.**
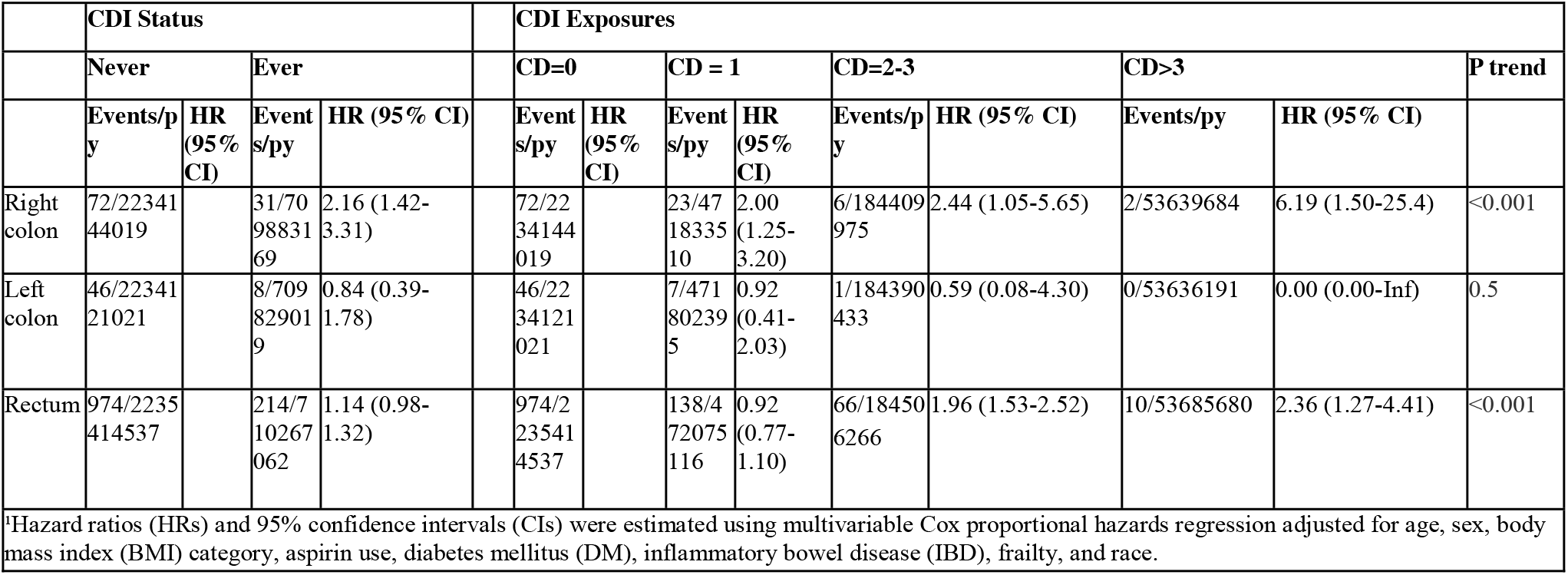
Associations of Any CDI and Dose-Dependent *Clostridioides difficile* Exposure With Colorectal Cancer Mortality by anatomical site (VHA, 2000–2025)

**Table 7.**
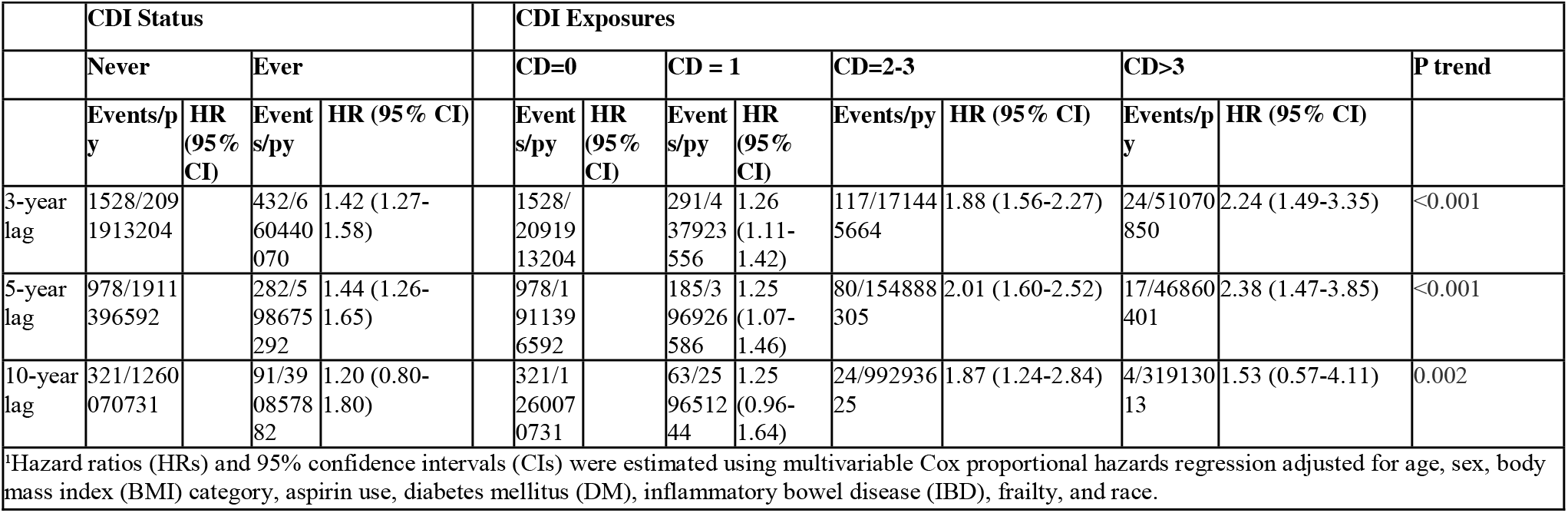
Associations of Any CDI and Dose-Dependent *Clostridioides difficile* Exposure With Colorectal Cancer Incidence by time lag (VHA, 2000–2025)

The observed dose-dependent risk was stronger at shorter lag times. At 3 years and 5 years, the dose-dependent pattern of more than one episode was associated with significantly increased risk for CRC incidence although it became weaker at 5 years and completely attenuated at 10 years. Similarly, the observed dose-dependent risk for mortality was strongest at 3 and 5 years but also attenuated at 10 years (see Table 4, Table 7).

## Discussion

In this nationwide, retrospective healthcare database cohort study, recurrent positive *C. difficile* testing was independently associated with increased CRC incidence and mortality in a dose-dependent manner, particularly among patients with IBD. This study further strengthens the epidemiological evidence linking repeated *C. difficile* test positivity to future CRC incidence and CRC mortality, and aligns with experimental mouse models demonstrating that persistent toxigenic *C. difficile* colonization promotes tumorigenesis by mediating pro-oncogenic inflammatory and colon epithelial signaling pathways.

This study extends and validates our prior multisite retrospective cohort analysis (12) using longitudinal medical record data from Michigan Medicine and Johns Hopkins Medicine, which demonstrated that repeated *C. difficile* test positivity was significantly associated with a two-fold increased risk of incident CRC, whereas a single episode was not associated. In this larger VHA retrospective cohort analysis, we confirmed these findings and further demonstrated a dose-dependent relationship, with progressively higher CRC risk associated with increasing CDI episodes (2–3 episodes and >3 episodes). Consistent with the prior study, no association was observed for individuals with only one CDI episode. Consistent with the mouse model data, these findings suggest the hypothesis that persistent *C. difficile* exposure rather than a single acute infection may underlie the results in the observed increase in CRC risk (10).

Prior studies investigating the relationship between CDI and CRC found conflicting results although they were limited by study design. A Florida Medicaid claims-based study observed that CDI diagnosis was associated with an increased risk for CRC development in the subsequent 4 years (13). However, this claims-based study relied on ICD billing codes rather than laboratory testing resulting in only 0.3% with a history of CDI. This is substantially lower than the 19% observed in our study population, however our study consisted of symptomatic patients undergoing CDI testing. CDI diagnosis based on claims data raises concern for misclassification bias and underestimation of *C. difficile* exposure. Conversely, in another national claims-based database study that matched patients with CDI to those without CDI, CDI exposure appeared protective against CRC except in obese patients, in whom CDI exposure increased the risk for CRC (14). However, no time lag was applied, allowing CDI and CRC diagnoses to occur concurrently. Failure to address temporality introduces substantial risk of reverse causality particularly because CRC is associated with microbiota dysbiosis, potentially creating a niche for *C. difficile* colonization. Furthermore, overmatching on many variables including age, sex, Charleson Comorbidity Index, CDI treatment, and obesity may have limited the analysis. Several of these matching variables such as sex and obesity may act as effect modifiers in the association between CDI exposure on CRC risk, while others may exist on the causal pathway. Another hospital-based retrospective study investigated the association between positive C. *difficile* testing and colorectal neoplasia identified on colonoscopy and pathology reports with outcomes lagged by five years (15). They did not find an association, although their study had limited statistical power due to a small sample size.

Importantly, our study also assessed the association between *C. difficile* test positivity and CRC mortality, which has never been explored before. Unlike CRC incidence, CRC specific mortality risk was elevated even among patients with a single positive *C. difficile* test and was stronger than that observed for CRC incidence. Similar to CRC incidence, however, it also demonstrated a dose-dependent relationship. Lag time analysis revealed that CRC incidence risk peaked at three years following initial toxigenic *C. difficile* exposure whereas CRC mortality risk peaked at five years. This suggests *C. difficile* presence may play a stronger role in CRC progression and tumor aggressiveness rather than CRC initiation. This finding aligns with the epidemiologic framework proposed by Welch et al. in which exposures that affect mortality more than incidence may influence disease lethality rather than initiation (18). Although the attenuation noted at longer lag times may also reflect reduced sample size at extended follow-up.

Consistent with this finding of a stronger magnitude of association observed for CRC mortality, prior studies have reported a higher prevalence of *C. difficile* colonization among patients with CRC, with the highest rates seen in advanced disease. In one study, 16% of CRC patients presenting for surgery had *C. difficile* strains isolated from stool samples and this was even more frequent (22%) among patients with lymph node metastases (19). These findings raise the possibility that persistent *C. difficile* colonization may be linked to tumor progression and more aggressive disease biology rather than simply reflecting increasing mortality through severe CDI. Adding further complexity, prior studies have also demonstrated higher all-cause mortality among cancer patients with CDI. A retrospective analysis of the NIS database (2016 to 2020) found that among patients with CDI, those with a concurrent diagnosis of CRC had an increased risk of all-cause in-hospital mortality (OR 2.24, 95% CI 1.13-4.44) (20). Similarly, a larger NIS analysis demonstrated that CDI was also independently associated with 62% increased odds of all-cause in-hospital mortality across all cancer types (9). However, other evidence suggests that mortality directly attributable to CDI itself may be relatively low in patients with cancer. In a multicenter study, only 4.9% of deaths were directly attributable to CDI despite an overall 90-day mortality of 22.3% (21) suggesting that excess mortality is driven by underlying malignancy rather than death from complicated CDI. Given our finding that a prior history of *C. difficile* exposure increased risk of CRC mortality, persistence of *C. difficile* colonization, particularly on the colon mucosa, may increase the risk of CRC progression and mortality.

Notably, both our prior study and this study identified evidence for effect modification by sex, although both analyses were limited by sample size. In the prior two-site cohort analysis, stratified analyses revealed a significant association between *C. difficile* exposure and CRC incidence among females (adjusted HR for females, 2.39 [95% CI, 1.34-4.26]) but not in males (adjusted HR for males, 1.73 [95% CI, 0.80-3.37]), however the interaction term was not statistically significant (p=0.10). In this current study, the association between *C. difficile* test positivity and CRC incidence and mortality was higher among women in the lower exposure risk categories and the interaction term was significant. However, the relatively small number of females in the VHA population resulted in fewer individuals in the higher exposure categories and limited CRC events precluding robust evaluation for dose-dependent association in the highest exposure category. Notably, there are many past studies showing an association between CDI outcome and sex (22).

We also found compelling evidence of effect modification by IBD status. Although both individuals with and without IBD demonstrated a dose-dependent association between *C. difficile* exposure and both CRC incidence and mortality, the magnitude of this association was greater in patients with IBD, particularly for CRC mortality. Notably, patients with IBD who experienced more than three positive *C. difficile* tests were more than three times as likely to die of CRC, compared to a 1.5 times increased risk among patients without IBD. This is in line with what we would have expected from known associations observed between CDI and IBD in terms of risk of infection and adverse IBD outcomes following CDI (23-25).

These findings are biologically plausible given emerging evidence that *C. difficile* may directly promote colorectal tumorigenesis and cancer progression through toxin-mediated inflammatory and epithelial signaling pathways. Drewes et al demonstrated that TcdB toxin promoted CRC through activation of oncogenic Wnt signaling, increased reactive oxygen species production, and induction of protumorigenic immune responses characterized by infiltration of activated myeloid cells and IL-17-producing lymphoid cells (10). These findings suggest chronic *C. difficile* exposure may contribute to CRC carcinogenesis through persistent toxin-mediated epithelial damage, oxidative stress, chronic inflammation, and immune dysregulation. However, persistence of *C. difficile* in human feces and/or mucosa has yet to be rigorously examined over time in individuals who appear to resolve symptomatic CDI or who develop rCDI.

These mechanisms may be amplified in patients with IBD. Severe or poorly controlled IBD is a known risk factor for CDI and emerging evidence suggests shared genetic polymorphisms that predispose to chronic mucosal inflammation may increase susceptibility to both IBD and CDI (26-28). Furthermore, experimental models support a bidirectional relationship in which IBD-associated inflammation promotes *C. difficile* colonization even in the absence of antibiotics. In turn, concurrent IBD and CDI results in synergistically worsened colonic mucosal inflammation (29), whereas treatment of inflammation in this mouse model shifted the microbiome toward a *C. difficile* colonization resistant state, strengthening the mediating role of inflammation in this pathway. Superimposed *C. difficile* toxin-mediated inflammation may therefore accelerate the established effects of IBD on chronic mucosal injury and microbiome dysregulation, further promoting dysplasia and colorectal carcinogenesis through exaggerated cytokine activation, oxidative stress, cell signaling, and DNA damage (30). This provides a biologically plausible explanation for our findings that association between CDI history and CRC incidence and mortality was strongest among patients with IBD.

This study has many strengths. This nationwide study using the VHA Corporate Data Warehouse represents the largest study to investigate the association between *C. difficile* test positivity and subsequent risk of CRC. The large sample size provided sufficient power to evaluate dose-dependent associations and to conduct subgroup analyses, including assessment of effect modification by sex and IBD status. The long duration of follow-up available within the VHA electronic health record allowed lag time sensitivity analyses, enabling us to identify the temporal window in which the associations are strongest. Like our prior two-hospital cohort study, a major strength of our approach was the use of *C. difficile* laboratory test results rather than ICD coding to define exposure, thereby reducing the risk of misclassification inherent in bill claims. Additionally, our decision to stratify exposure by nominal dose (single versus multiple positive tests) enabled a higher-resolution assessment of chronic or recurrent toxigenic *C. difficile* exposure without reliance on ICD codes for recurrent CDI. The use of time-varying exposure models with cumulative dose updates further allowed us to fully leverage longitudinal data and improve estimate precision.

This study has limitations inherent to any medical records-based retrospective study. Information on behavioral risk factors including alcohol use, diet, and exercise was unavailable. The study population was limited to veterans receiving their care within the VHA, which limits the data generalizability given the underrepresentation of women and certain racial groups in this patient population. Although the total sample size enables subgroup analyses, smaller subgroup sizes may have limited statistical power to detect differences in the outcome by sex, race or anatomic subsite. More representative datasets are needed to further assess racial, sex and anatomic subgroup differences. Finally, we did not adjust for or exclude individuals with hereditary colon cancer because there is no specific billing code to identify Lynch syndrome.

## Conclusion

These findings extend animal model evidence, epidemiologically establishing *C. difficile* exposure as an independent risk factor for subsequent colorectal tumorigenesis and supporting investigation into recurrent CDI, especially among patients with IBD as a potential modifiable CRC risk factor. Future work will include validation of our findings using non-VA healthcare systems that will enhance the generalizability of our findings. Translational studies are needed to integrate microbiome/microbiota community studies within prospective cohorts of patients with CDI and colorectal polyps. Such investigations will enable assessment of longitudinal *C. difficile* persistence and microbiome composition that may mediate the relationship between *C. difficile* exposure and colorectal neoplasia. Our findings have implications for evaluation of CRC risk prediction models and whether recurrent CDI constitutes a high-risk group who may benefit from modified CRC prevention screening and surveillance strategies. Ultimately, if CDI does accelerate CRC pathogenesis, a key question will be whether microbiome-modifying CDI treatments (e.g., Rebyota and Vowst) not only reduce CDI recurrence but also lower future risk of CRC.

## Author contributions

<u>Conceptualization</u>: S.R., S.M.A., N.O.M.,K.R., C.L.S.; <u>Methodology</u>: S.R, S.M.A., O.W.,K.R., M.J.S., C.L.S. <u>Formal Analysis</u>: S.R., O.W, M.J.S, K.R;. <u>Investigation</u>: S.R., S.M.A., O.W., K.R., C.L.S., <u>Resources</u>: S.R., N.O.M, K.R., C.L.S, <u>Writing – Original Draft:</u> S.R.; <u>Writing – Review & Editing</u>: all authors; <u>Visualization</u>: S.R., <u>Supervision</u>: C.L.S., K.R.; <u>Funding Acquisition</u>: S.R., C.L.S., M.J.S.

## Data Availability

The data generated in this study are available within the article and its supplementary data files.

## Abbreviations

CRC: Colorectal Cancer
CDI: *Clostridioides difficile* Infection
rCDI: Recurrent *Clostridioides difficile* Infection
TcdB: *C. difficile* Toxin B
ICD: International Classification of Disease
CD: *C. difficile*
IBD: Inflammatory Bowel Disease
95% CI: 95% Confidence Interval
HR: Hazard Ratio

